# Downward bias in the association between APOE and Alzheimer’s Disease using prevalent and by-proxy disease sampling in the All of Us Research Program

**DOI:** 10.1101/2025.05.22.25328175

**Authors:** Clayton O. Mansel, Valentina Ghisays, Jonathan D. Mahnken, Russell H. Swerdlow, Eric M. Reiman, Jason H. Karnes, Joshua C. Denny, Olivia J. Veatch

## Abstract

**Background:** Recent genome-wide association studies (GWAS) for Alzheimer’s Disease and related dementias (ADRD) have increased statistical power via larger analysis datasets from biobanks by 1) including non-age-matched controls and prevalent cases, and/or 2) including individuals who report a family history of ADRD as proxy cases. However, these methods have the potential to increase noise and distort genetic associations which are important for genomic-informed prevention and treatment of ADRD. Here, we sought to understand how the effect sizes of genetic associations in ADRD could be sensitive to these methodological choices, using *APOE* genotypes as an example.

**Methods:** Participants in the *All of Us* Research Program over the age of 49 at enrollment (n=258,693) were assigned one of four categories: incident ADRD (developed after enrollment in *All of Us*), prevalent ADRD (present on enrollment), proxy ADRD (participant noted a family history of ADRD), and control (no history or diagnosis of ADRD). Dementia diagnoses were determined using available Electronic Health Records (EHR) and *APOE* genotype was determined using whole-genome sequencing. Effect sizes for the associations between *APOE* risk alleles and ADRD diagnoses were compared using polychotomous logistic regression.

**Results:** The mean age of the cohort was 67±10 years, and it was 58% female; 63% clustered predominantly with European genetic reference populations. Among the participants, 3,107 (1.2%) had prevalent ADRD, 301 (0.1%) had incident ADRD, and 19,910 (7.7%) reported a family history of ADRD (proxy ADRD). Both prevalent and proxy ADRD had attenuated associations with *APOE* genotype compared to incident ADRD. The adjusted generalized ratio (95% CI) (AGR) for incident ADRD for *APOE* ε4 heterozygotes was 2.95 (2.31-3.74) compared to 2.10 (1.96-2.24) and 1.42 (1.32-1.55) for proxy and prevalent ADRD, respectively. For *APOE* ε4 homozygotes, the effect sizes were even more different. Furthermore, *APOE* association effect sizes increased when restricting the control (no ADRD) group to older age brackets.

**Conclusions:** Our study highlights how genetic associations with ADRD can be sensitive to how cases are defined in biobanks like *All of Us*, with effect sizes downwardly biased when using prevalent or by-proxy cases compared to incident cases.

## Background

Alzheimer’s Disease and related dementias (ADRD) are a devastating class of progressive neurodegenerative diseases of which over 7 million are affected in the U.S. alone and there is no cure [1,2]. AD in particular has a long prodromal preclinical phase in which biomarkers such as glucose hypometabolism and amyloid-beta plaques appear in the brain years before noticeable cognitive decline [3]. This has led to efforts for early detection and prevention, including genomic-informed risk assessments given that ADRD is highly heritable [4]. These genomic-informed risk assessments rely on inferences drawn largely from case-control and observational study designs and are thus subject to well-documented selection and survival bias [5–7]. In an effort to increase sample size and thus statistical power, recent genome-wide association studies (GWAS) for ADRD have incorporated observational data from large biobanks such as the UK Biobank and *All of Us* Research Program [8,9]. This has led to controversies including the use of so-called “proxy cases” in which an individual with a self-reported family history of ADRD in a first degree relative, but no evidence of cognitive decline, is included as a proxy case [10]. For example, in a recent GWAS for ADRD, 49,275 out of 111,326 total cases were proxy cases from the UK Biobank [8].

The proportion of individuals in modern ADRD GWAS that come from biobanks is quickly dwarfing ADRD-specific cohorts, despite their potential biases [5,11,12] and challenges with identifying clinical ADRD cases using electronic healthcare records (EHR) [13]. When an individual enrolls in a biobank such as the UK Biobank, he or she often consents to adding years of previous EHR records [14], including potential diagnoses of dementia. Because individuals cannot have cognitive impairment when they give informed consent, this can lead to a cohort of “prevalent cases” (i.e., cases diagnosed prior to enrollment in a biobank) that are healthier or decline slower than the general population. In addition to clinical cases, biobank proxy cases can be plagued by non-participation bias in optional family health history questionnaires—which have been shown to have genetic predictors of participation, including variants associated with ADRD [15]. Previous studies have shown downward bias in the effect size estimates for ADRD risk alleles when using proxy cases in the UK Biobank [16,17]. We note that the UK Biobank has been shown to be healthier than the general UK population [18] while participants in the *All of Us* Research Program have higher disease prevalence compared to the general U.S. population [19]. It is currently unknown if this alleviates the downward bias of prevalent and by-proxy disease sampling shown in previous studies in the UK Biobank.

Among the most consistently replicated GWAS loci for ADRD is the *APOE* genotype which is a strong predictor of dementia liability, currently explaining more phenotypic variability than polygenic scores constructed from hundreds of small-effect loci derived from GWAS [4,20,21]. Individuals with one copy of the *APOE* ε4 allele have a 3-4 fold increases risk for AD while two copies of ε4 carries a 12-15 fold increased risk [22,23]. As such, *APOE* genotyping is currently recommended as a part of evidence-based dementia prevention [24] and is likely to be a part of genomic-informed dementia care prior to polygenic scores which require much more optimization [21,25,26]. Given these previous studies, we sought to understand how genetic associations in ADRD could change depending on the definition of dementia, using *APOE* as an example for an effect likely present in many genes. We leverage the *All of Us* Research Program, a multisite, U.S.-based biobank, including many individuals from backgrounds whose health outcomes have not been extensively studied in biomedical research [27]. Our study provides early insight into whether the biases of proxy cases and incident vs prevalent disease sampling is present in one of the most populous and diverse biobanks in the world.

## Methods

### *All of Us* and Inclusion Criteria

The *All of Us* Research Program is a longitudinal, multi-site biorepository of participants’ health data including surveys, EHR, and other data linked to genomic data including whole-genome sequencing (WGS) [27,28]. It began enrollment nationally in 2018. EHR data from dozens of healthcare providers across the U.S. are harmonized using the Observational Medical Outcomes Partnership (OMOP) Common Data Model [29]. The data were acquired under the Data Use and Registration Agreement between the University of Kansas Medical Center and *All of Us*. As of March 2025, 633,540 participants had completed initial consent and enrollment for the Curated Data Repository v8. Participants were included in the present study if they 1) had available EHR data 2) had WGS data for *APOE* genotyping 3) were assigned male or female at birth and 4) were over the age of 49 at enrollment (enrollment date ranges from 05-31-2017 to 09-14-2023). The total sample size meeting these criteria was 258,693 participants.

### Definition of Phenotypes

For incident and prevalent dementia, a previously validated computable phenotype was used based on five or more visits with matching ICD-9/ICD-10 codes or one visit with a matching drug prescription [30]. Individuals meeting these criteria fully (i.e., recording their fifth visit) prior to their *All of Us* enrollment date were labelled as prevalent dementia while individuals meeting criteria after enrollment were labelled as incident dementia. For proxy dementia, the Personal and Family Health History survey was used which was filled out by (26.5%) of the participants included in this study. If the individual answered “mother”, “father” or “sibling” to the question “Including yourself, who in your family has had dementia (includes Alzheimer’s, vascular, etc.)?” they were labelled as proxy dementia. All remaining individuals who did not meet criteria for incident, prevalent, or proxy dementia were labelled control. Participants’ age was defined as age at enrollment in *All of Us*. Sex assigned at birth was self-reported. Global genetic ancestry reference population similarity was provided by *All of Us* for each participant and was inferred using a random forest classifier trained on samples from 1,000 Genomes and Human Genome Diversity Project samples [28]. We assign individuals to genetically defined ancestry groups based on their similarity to global reference populations, with the understanding that these categories are imperfect proxies for the continuous and admixed nature of human genetic variation. These groupings are used solely to enable analyses of genetic variation across diverse populations and are not intended to represent social or racial identities. EHR length was determined by the difference between the earliest and latest visit date across multiple OMOP Common Data Model tables.

### *APOE* genotyping

*All of Us* provides qualified researchers access to WGS data that has undergone quality control procedures outlined extensively elsewhere [28]. This study specifically used the Allele Count/Allele Frequency (ACAF) unphased callset comprised of single nucleotide polymorphisms (SNPs) meeting a population-specific allele frequency threshold of >0.01 or allele count >100. *APOE* SNPs were extracted using *PLINK* 1.9 and *APOE* genotypes were determined based on the combined genotypes at rs429358 and rs7412 [31] and imputed using R as previously described [32]. Because multiple phenotype groups (e.g., incident dementia) contained *APOE* genotype counts <20, we binned *APOE* genotypes into three groups: *APOE* ε4 non-carrier (ε2/ ε2, ε2/ ε3, ε3/ ε3), *APOE* ε4 heterozygote (ε2/ ε4, ε3/ ε4), and *APOE* ε4 homozygote (ε4/ ε4).

### Statistical Analysis

Summary statistics compared the mean (standard deviation) for continuous variables and n (%) for categorical variables. Group level comparisons were performed via Pearson chi-square tests for categorical variables and one-way analyses of variance for continuous variables. To determine the association between *APOE* genotype and dementia status, we modeled the nominal response variable dementia status using polychotomous logistic regression with a generalized logit function [33]. We included age at enrollment, sex assigned at birth, and global genetic ancestry as covariates considering extensive evidence of these variables interacting with the *APOE* genotype to influence risk for AD [22,34–36]. The results are presented as adjusted generalized ratios (AGRs) which can be interpreted as ratio of the likelihoods of each definition of dementia compared to control which was the referent group. The AGRs for *APOE* genotype and proxy dementia were adjusted by a factor of 2 to account for the genetic relatedness between parent and child. This adjustment has been shown to be accurate for case/control odds ratios for alleles with small to medium effect sizes [10]. To ensure that this adjustment appropriate for our study, we compared adjusting the proxy AGR by a factor of 2 with another method derived directly from the allele frequencies in the cases and controls as shown in Liu et al., 2017 [10] and found that the adjusted AGR is similar (Supplementary Table 1).

Recent ADRD GWAS have stopped using age-matched controls. For example, the most recent large GWAS had controls with a mean age at assessment of 57.9 compared to 67.2 for the cases [8]. Given this, we also ran our analyses using different age cut-offs in the control group (e.g., >49yrs, >59yrs, >69yrs, >79yrs). All analyses were performed in R v4.4.0 and RStudio v2024.04.0.

## Results

Of the 245,693 individuals included in the analysis dataset, 301 (0.1%) had incident dementia, 3,107 (1.3%) had prevalent dementia and 19,910 (8.1%) had proxy dementia (Table 1). The overall mean (standard deviation) EHR length was 10.97 (9.60) years. Across controls and dementia subgroups, all were majority (>50%) assigned female at birth with the proxy dementia subgroup at a notably higher proportion (64.7%) (Table 1). The mean age of the subgroups ranged from 66.5 to 79.2 years. The subgroup with the highest mean age was incident dementia (79.3) followed by prevalent dementia (74.2), proxy dementia (68.6) and then controls (66.5) (Table 1). The proportion of participants carrying 1 *APOE* ε4 allele varied widely from 24.3% of the control group to 41.2% of the incident dementia subgroup (Table 1). The incident dementia subgroup also had the highest proportion of *APOE* ε4 homozygotes (7.0%) compared to the other groups (Table 1). All subgroups were mostly comprised of individuals with European ancestry. The control group had the smallest proportion of European ancestry at 61.0% followed by incident dementia at 68.1%, prevalent dementia at 69.9%, and proxy dementia with 85.1% of individuals (Table 1).

**Table 1.** Age, Sex, *APOE* genotype, and Genetic Ancestry for 258,693 *All of Us* participants.

|  | Control | Incident Dementia | Prevalent Dementia | Proxy Dementia | p value |
| --- | --- | --- | --- | --- | --- |
| n (%) | 235375 | 301 | 3107 | 19910 |  |
| Demographics |  |  |  |  |  |
| Female Sex (%) | 135004 (57.4) | 158 (52.5) | 1597 (51.4) | 12882 (64.7) | <0.001 |
| Age (mean [sd]) | 66.5 (10.0) | 79.3 (8.2) | 74.2 (10.5) | 68.8 (8.4) | <0.001 |
| *APOE* Genotype |  |  |  |  |  |
| *APOE* ε4 non-Carrier | 173267 (73.6) | 156 (51.8) | 2091 (67.3) | 13491 (67.8) | <0.001 |
| *APOE* ε4 Heterozygote | 57128 (24.3) | 124 (41.2) | 873 (28.1) | 5849 (29.4) |  |
| *APOE* ε4 Homozygote | 4980 (2.1) | 21 (7.0) | 143 (4.6) | 570 (2.9) |  |
| Global Genetic Ancestry |  |  |  |  |  |
| African | 50340 (21.4) | 50 (16.6) | 441 (14.2) | 1372 (6.9) | <0.001 |
| American Admixed/Latino | 34978 (14.9) | 40 (13.3) | 438 (14.1) | 1211 (6.1) |  |
| East Asian | 4112 (1.7) | ** | 34 (1.1) | 262 (1.3) |  |
| European | 143471 (61.0) | 205 (68.1) | 2161 (69.6) | 16945 (85.1) |  |
| Middle Eastern | 621 (0.3) | ** | ** | 39 (0.2) |  |
| South Asian | 1853 (0.8) | ** | ** | 81 (0.4) |  |

Total sample size is 245,693 participants in the *All of Us* Research Program who identified as male or female at birth and were over the age of 49 at enrollment. Incident and prevalent dementia defined based on whether the participant met diagnostic criteria before or after they enrolled in *All of Us* (see methods). Proxy dementia defined based on whether the participant reported a history of dementia in a first-degree relative. Categorical variables are presented as n (%) and continuous variables presented as mean (standard deviation). p values are unadjusted and correspond to $\chi^2$ tests for categorical variables and one-way analyses of variance (ANOVA) for continuous variables. \*\*Participant counts <20 suppressed in accordance with *All of Us*'s Data and Statistics Dissemination Policy

The relative ratio of the proportion belonging to each dementia status group divided by the proportion of controls among *APOE* ε4 heterozygotes (relative to non-carriers) and homozygotes (relative to non-carriers) is shown in Figure 1 and Supplementary Table 2 and is presented as adjusted generalized ratios (AGRs) (see Methods). For the *APOE* ε4 heterozygotes, the adjusted generalized ratio (AGR) (95% CI) was highest for incident dementia at 2.95 (2.31-3.74). In comparison, both prevalent and proxy dementia were less associated with AGRs of 1.43 (1.32-1.55) and 2.10 (1.96-2.24), respectively (Figure 1; Supplementary Table 2). For *APOE* ε4 homozygotes, again the AGR was highest for incident dementia at 7.23 (4.55-11.54); for prevalent dementia the AGR was 3.13 (2.63-3.73); for proxy dementia the AGR was 3.51 (2.93-4.21) (Figure 1; Supplementary Table 2). We note that in both *APOE* ε4 heterozygotes and homozygotes, the prevalent and proxy dementia AGR was significantly lower than the incident dementia AGR (95% confidence interval limits).

**Figure 1:**
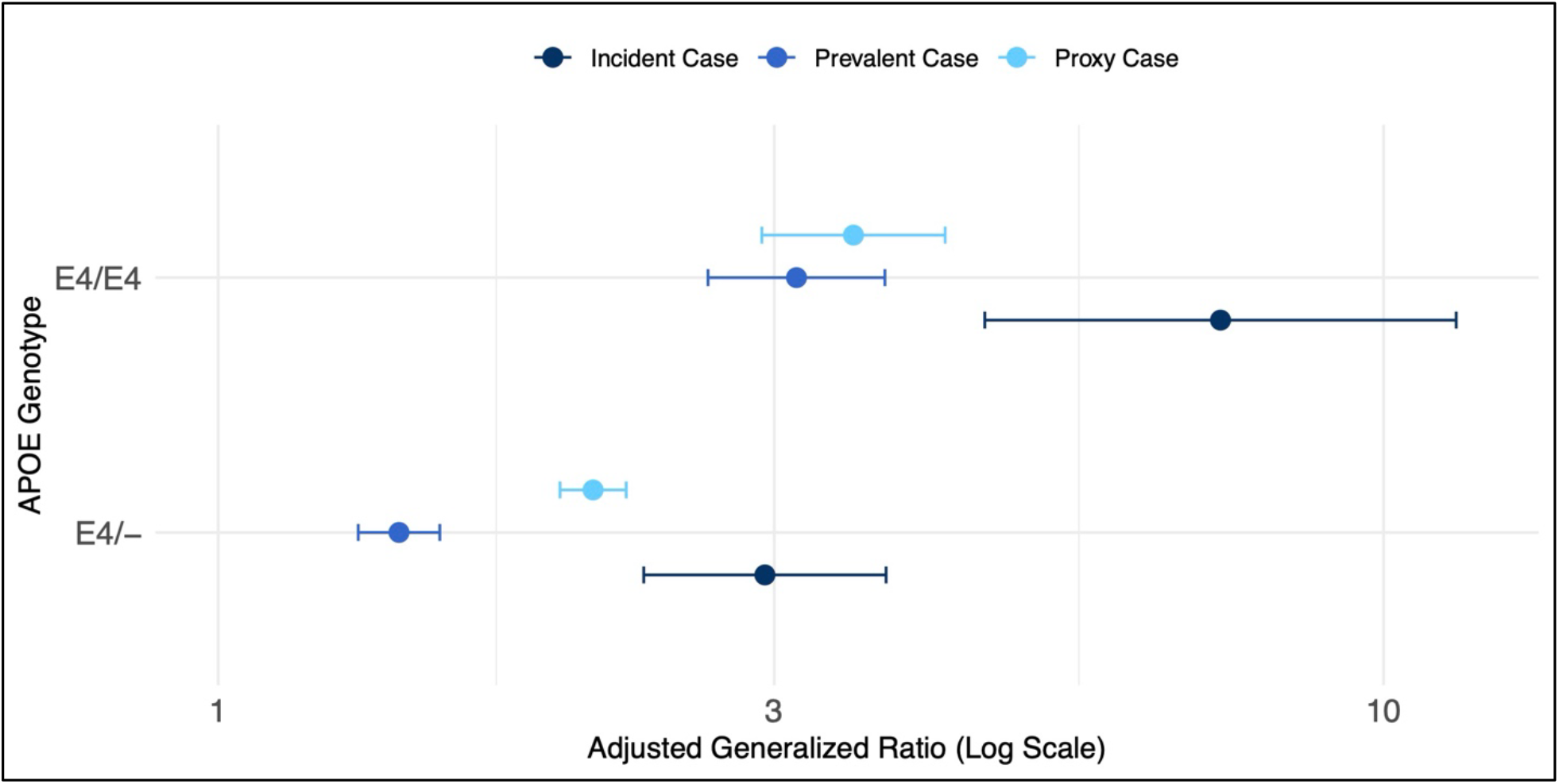
Adjusted generalized ratios for the association between *APOE* ε4 genotype and dementia status in *All of Us*. Total sample size is 245,693 participants in the *All of Us* Research Program who identified as male or female at birth and were over the age of 49 at enrollment. Incident and prevalent dementia defined based on whether the participant met diagnostic criteria before or after they enrolled in *All of Us* (see Methods). Proxy dementia defined based on whether the participant reported a history of dementia in a first-degree relative. Adjusted generalized ratio’s plotted (log scale) and derived from a polychotomous logistic regression with dementia status as the outcome (outcome referent = control) adjusted for age at enrollment, sex assigned at birth, and global genetic ancestry. Proxy dementia AGRs were adjusted by a factor of 2 to account for the genetic relatedness between the parent and child.

To account for the fact that some control participants may be mislabeled because of a short history of EHR visits, we restricted our dataset to only individuals with greater than one year of EHR visits (n = 218,542) and found that this did not affect the association between *APOE* and dementia status (Supplementary Figure 1). We also restricted our dataset to just individuals who completed the Family Health History survey (n = 68,643) to prevent misattribution of control individuals who should be labelled as proxy dementia (Supplementary Figure 2). We found that, again, both proxy and prevalent dementia AGRs were significantly smaller compared to incident dementia. One of the purported benefits of using proxy disease sampling is to help alleviate misclassification bias from controls who are too young to receive a diagnosis of ADRD. To address this, we reanalyzed the association between *APOE* and dementia status using different age cut-offs in the control group (e.g., >49yrs, >59yrs, >69yrs, >79yrs) and showed that, even for proxy cases, the older control groups numerically increased the AGRs (Figure 2).

**Figure 2:**
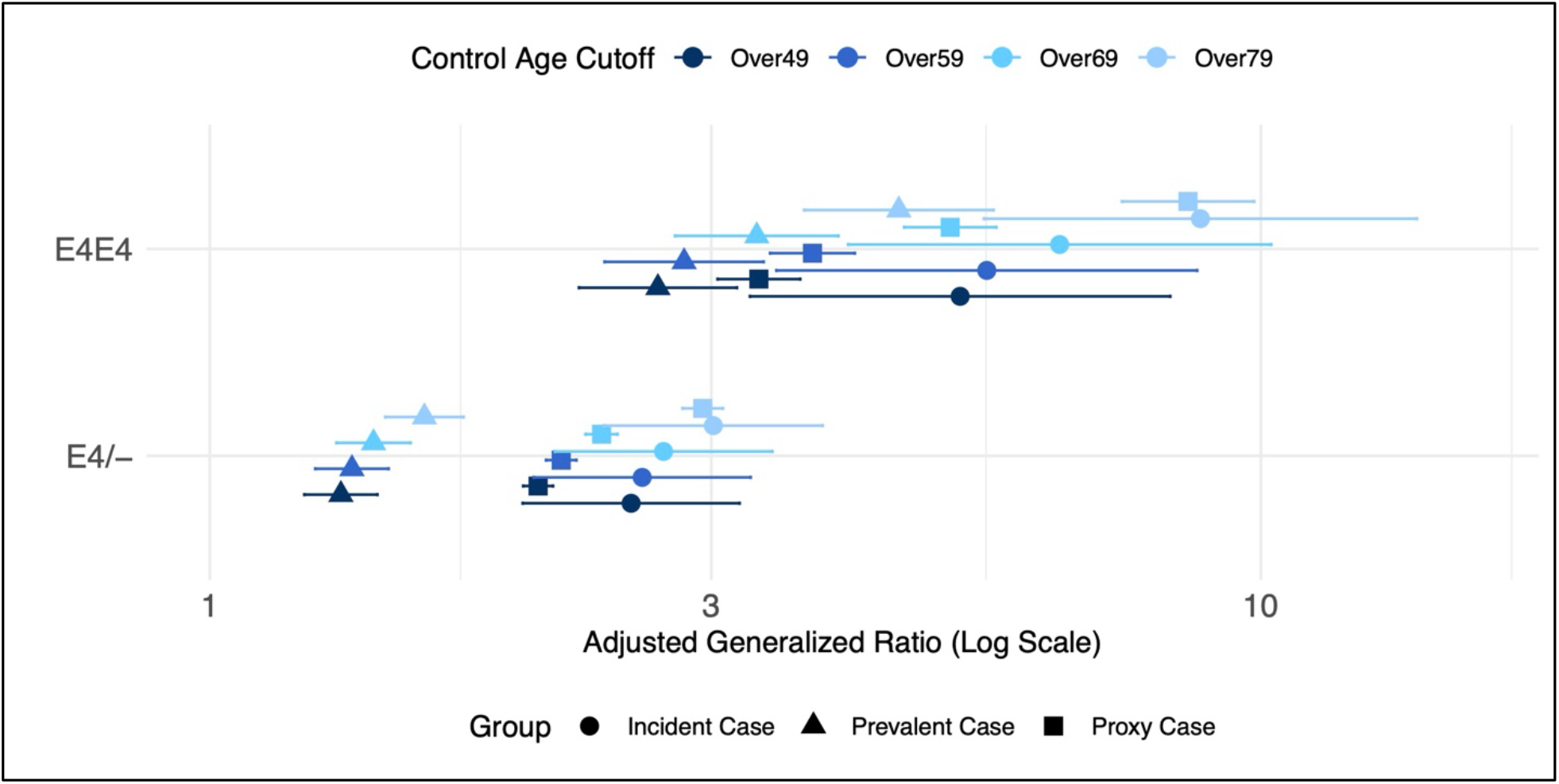
Adjusted generalized ratios for the association between *APOE* ε4 genotype and dementia status in *All of Us*. Total sample size is 245,693 participants in the *All of Us* Research Program who identified as male or female at birth and were over the age of 49 at enrollment. Incident and prevalent dementia defined based on whether the participant met diagnostic criteria before or after they enrolled in *All of Us* (see methods). Proxy dementia defined based on whether the participant reported a history of dementia in a first-degree relative. Adjusted generalized ratio’s plotted (log scale) and derived from a polychotomous logistic regression with dementia status as the outcome (outcome referent = control) adjusted for age at enrollment, sex assigned at birth, and global genetic ancestry. Data are stratified to include only non-demented participants over the age of 49 yrs (n = 258,693), 59 yrs (n = 192,438), 69 yrs (n= 112,215), and 79 yrs (n=48,352).

## Discussion

Our study demonstrates that using proxy and prevalent case definitions of ADRD significantly attenuates the observed association between *APOE* genotypes and disease risk when compared to incident cases. Proxy case definitions based on a family history of ADRD is a common approach in modern ADRD GWAS and other case/control study designs [8,37–39]. We demonstrated significant downward bias in genetic associations with proxy ADRD in *All of Us*, a finding corroborated by previous studies in the UK Biobank [16,17,40]. Similarly, we showed that prevalent ADRD cases in *All of Us* are less associated with *APOE* compared to incident cases. While biobanks like *All of Us* can take advantage of decades of EHR data on enrolled participants, our study highlights how this can underestimate the association of genetic risk factors like *APOE* in ADRD, especially when evaluating prevalent conditions whose presence may be negatively correlated with the probability of enrolling. Although AGRs, odds ratios (OR), and relative risk are not directly comparable, our estimate for the association between *APOE* and incident ADRD cases (AGR: 2.95 for ε4/- and 7.23 for ε4/ε4) appears closer to epidemiologic evidence estimating 3-4 fold increased risk for *APOE* ε4 heterozygotes and 12-15 fold increased risk for *APOE* ε4 homozygotes [22,23]. Furthermore, we showed that all effect sizes are greater when filtering the control group to older cohorts. This highlights a major challenge for ADRD case-control study designs in which controls can be incorrectly labelled given that the disease starts long before cognitive symptoms appear. Modern blood-based biomarkers could help remedy this issue by at least excluding controls who are positive for beta amyloid in the brain [41].

For the purposes of discovering novel biological pathways to prioritize for further functional validation, the use of biobanks and proxy cases in GWAS may be sensible [10]. But for genomic-informed risk assessments, in which the magnitude of the association between genotype and phenotype affects the potential absolute risk reduction of costly interventions [42], we argue that more precision is needed. Proponents of proxy case genetic association studies (GWAX) argue that the effect sizes are similar between GWAS and GWAX for large-effect loci [8,10] and that there is not significant heterogeneity of individual single nucleotide polymorphism (SNP) effect sizes [17]. However, using mendelian randomization, others have found significant genetic heterogeneity when using clinical ADRD vs proxy ADRD [40]. A recent GWAS found that only 0.6% of loci identified using proxy cases from *All of Us* were present when using clinical AD cases—including many with the opposite direction of effect [9]. Our study demonstrated that prevalent cases in *All of Us* have a downward bias in genetic associations with ADRD to a similar magnitude as proxy cases. Thus, both sampling strategies may be inappropriate for building genomic-informed risk assessments.

Given the decreasing cost of genotyping and genome sequencing, genomic-informed medicine presents an enticing opportunity to improve risk stratification and tailor treatments for many diseases. For instance, previous work in atherosclerotic cardiovascular disease has proven the utility of genomic-informed risk assessments which have been shown to successfully motivate behavior-change in almost half of a recent prospective cohort [43]. The same potential exists for dementia prevention [44]. However, dementia faces unique challenges in genetic association studies given that it is a disease of late life, has a long prodromal phase, and has no validated biomarker that is widely used for accurate diagnosis outside of specialist memory centers. GWAS including proxy cases and prevalent biobank cases may be distorting important genetic associations needed to build effective genomic-informed prediction models and thus may be doing more harm than good. The inferences drawn from GWAS matter a lot to this effort as evidenced by a recent benchmarking study which found that the choice of GWAS had the greatest effect of any methodological choice in genomic-informed dementia prediction modelling [45].

There are potential remedies to the biases demonstrated in this study. Future genetic association studies could incorporate survival data using time-matched sampling or weighting by the time observed between diagnosis and sampling [46,47]. Second, if the probability of being enrolled in the biobank could be estimated, then inverse probability weighting could be used to generate less biased estimates [48]. Third, each passing year provides more and more incident cases in biobanks, remedying this issue over time. However, any statistical correction of bias will pale in comparison to resolving the issue at the GWAS itself. Modern ADRD GWAS have larger and larger sample sizes but smaller and smaller single nucleotide polymorphism (SNP) heritability estimates [49]. We argue that GWAS could be tailored to the specific goal in mind.

For genomic medicine inferences, GWAS should not use proxy cases, use age-matched controls, use incident cases to minimize selection biases where possible, and, given the available of accurate biomarkers now for ADRD [41], recommend use of biomarkers where possible to minimize misclassification of case-control status. At the very least, the summary statistics for associations excluding proxy cases should be made available to the research community, something that was lacking in the most recent large ADRD GWAS [8].

Our study was limited to the use of EHRs to infer ADRD case/control status, which is not as accurate as diagnoses at specialist memory centers and prevented us from distinguishing between AD and other closely related dementias which could lead to an underestimation of *APOE*’s association with disease. We were also not able to evaluate *APOE* ε2 allele-specific effects due to small sample sizes. In addition, we did not have a large enough sample size to stratify by race/ethnicity and thus could not determine whether the prevalent and proxy bias differed in non-White or Hispanic individuals. Notably, we had a much higher proportion of individuals of European ancestry in the proxy case cohort. We hypothesize that this could be because individuals identifying as White were more likely to fill out the Family Health History survey. Despite these limitations, our study highlights a fundamental question in genomic-informed medicine: how do we infer an association between a gene and a disease for use in the clinic? Our study showed how these associations are sensitive to changes in methodology. Early personalized interventions for ADRD are likely to be expensive and laborious [24], thus there exists a need for greater precision to determine who is likely to benefit the most.

## Data Availability

All data used in this study can be accessed by qualified researchers at allofus.nih.gov. Source code for these analyses is available within the *All of Us* Researcher Workbench at (https://workbench.researchallofus.org/workspaces/aou-rw-55fcf7f4/alzheimersdiseaseapoeprevalencevsincidencev8/data).

## Acknowledgements

We acknowledge *All of Us* participants for their contributions, without whom this research would not have been possible. We also thank the National Institutes of Health’s [All of Us Research Program](https://allofus.nih.gov/) for making available the participant and genetic data examined in this study. The All of Us Research Program is supported by the National Institutes of Health, Office of the Director: Regional Medical Centers: 1 OT2 OD026549; 1 OT2 OD026554; 1 OT2 OD026557; 1 OT2 OD026556; 1 OT2 OD026550; 1 OT2 OD 026552; 1 OT2 OD026553; 1 OT2 OD026548; 1OT2 OD026551; 1 OT2 OD026555; IAA #: AOD 16037; Federally Qualified Health Centers: HHSN 263201600085U; Data and Research Center: 5 U2C OD023196; Biobank: 1 U24 OD023121; The Participant Center: U24 OD023176; Participant Technology Systems Center: 1 U24 OD023163; Communications and Engagement: 3 OT2 OD023205; 3 OT2 OD023206; and Community Partners: 1 OT2 OD025277; 3 OT2 OD025315; 1 OT2 OD025337; 1 OT2 OD025276. In addition, the *All of Us* Research Program would not be possible without the partnership of its participants.

