## Supplementary Material for "Downward bias in the association between APOE and Alzheimer’s Disease using prevalent and by-proxy disease sampling in the All of Us Research Program"

**Supplemental Table 1. The association between *APOE* ε4 genotype and dementia diagnosis adjusted using two different methods**

| Method | Generalized Ratio (95% CI) for Proxy Dementia |
|---|---|
| Unadjusted | 1.31 (1.27-1.36) |
| Calculated directly using allele frequencies in cases/controls with equation from Liu et al., 2017 | 1.59 (1.51-1.66) |
| Adjusted by a factor of 2 | 1.71 (1.62-1.84) |

The association between *APOE* ε4 genotype and dementia diagnosis was tested via a polychotomous logistic regression with dementia status (control, incident, prevalent, and proxy dementia) as the outcome and no other covariates. To account for the genetic relatedness between parent and child, we adjusted the generalized ratio for proxy dementia using two different methods. First, we calculated the adjusted generalized ratio directly using the frequency of the *APOE* ε4 allele in the cases and controls and the equation in Liu et al., 2017 [10]. Then, we adjusted the generalized ratio by a factor of two as is recommended by [10] for small to medium effect size alleles and was done in the latest ADRD GWAS [8].

**Supplemental Table 2. The association between *APOE* genotype and incident, prevalent, and proxy dementia diagnosis in *All of Us***

| Dementia Status | Incident Dementia | Prevalent Dementia | Proxy Dementia |
|---|---|---|---|
| n | 301 | 3107 | 19910 |
| | Adjusted Generalized Ratio^ (95% CI) | | |
| *APOE* Genotype | | | |
| E4 Heterozygote | 2.95* (2.31-3.74) | 1.43* (1.32-1.55) | 2.10* (1.96-2.24) |
| E4 Homozygote | 7.23* (4.55-11.54) | 3.13* (2.63-3.73) | 3.51* (2.93-4.21) |

Total sample size is 245,693 participants in the *All of Us* Research Program who identified as male or female at birth and were over the age of 49 at enrollment. Incident and prevalent dementia defined based on whether the participant met diagnostic criteria before or after they enrolled in *All of Us* (see methods). Proxy dementia defined based on whether the participant reported a history of dementia in a first-degree relative. The association between *APOE* genotype and dementia diagnosis was tested via a polychotomous logistic regression with dementia status as the outcome. The referent group for dementia status was not demented (n=235,375). Proxy dementia AGRs were adjusted by a factor of 2 to account for the genetic relatedness between the parent and child. ^Adjusted generalized ratio's (95% Wald Confidence Intervals) presented for *APOE* ε4 heterozygotes and homozygotes adjusted for age at enrollment, sex assigned at birth, and global genetic ancestry. *p < 0.05 unadjusted (95% Wald Confidence Limits)

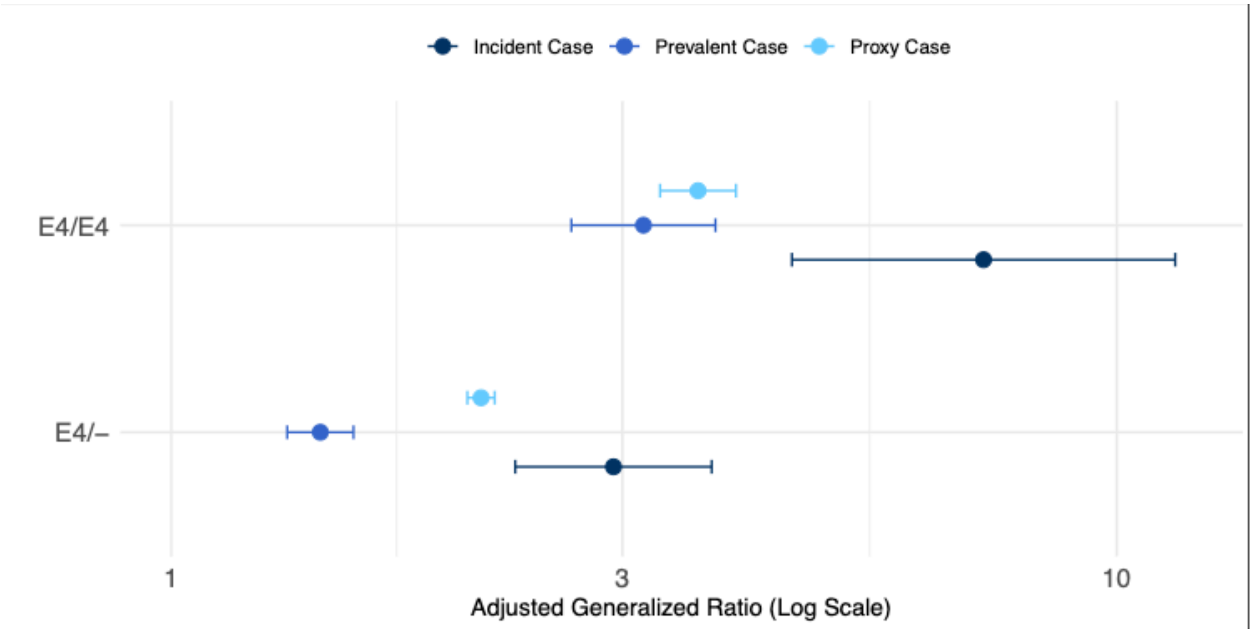


**Supplementary Figure 1: Adjusted generalized ratios for the association between *APOE* ε4 genotype and dementia status in *All of Us* in participants with > 1 year of EHR visits.** Total sample size is 218,542 participants in the *All of Us* Research Program who identified as male or female at birth, were over the age of 49 at enrollment, and had > 1 year of EHR visits. Incident and prevalent dementia defined based on whether the participant met diagnostic criteria before or after they enrolled in *All of Us* (see methods). Proxy dementia defined based on whether the participant reported a history of dementia in a first-degree relative. Adjusted generalized ratio's plotted (log scale) and derived from a polychotomous logistic regression with dementia status as the outcome (outcome referent = control) adjusted for age at enrollment, sex assigned at birth, and global genetic ancestry.

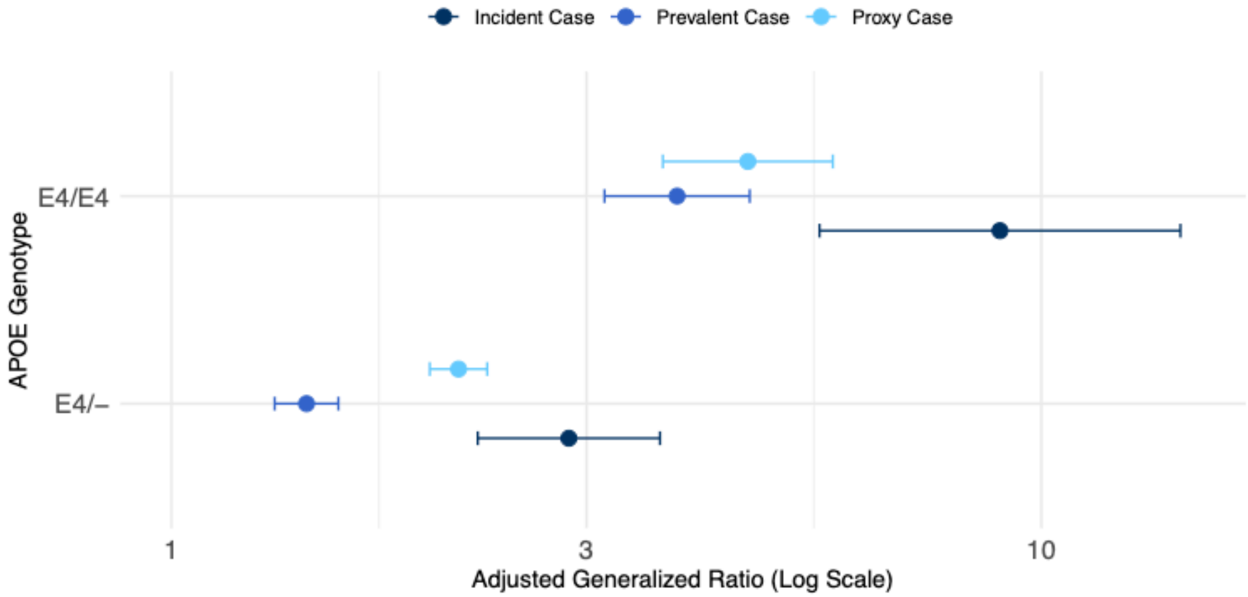


**Supplementary Figure 2: Adjusted generalized ratios for the association between *APOE* ε4 genotype and dementia status in *All of Us* in participants who filled out the Family Health History survey.** Total sample size is 68,634 participants in the *All of Us* Research Program who identified as male or female at birth, were over the age of 49 at enrollment, and filled out the Family Health History survey. Incident and prevalent dementia defined based on whether the participant met diagnostic criteria before or after they enrolled in *All of Us* (see methods). Proxy dementia defined based on whether the participant reported a history of dementia in a first-degree relative. Adjusted generalized ratio's plotted (log scale) and derived from a polychotomous logistic regression with dementia status as the outcome (outcome referent = Not Demented) adjusted for age at enrollment, sex assigned at birth, and global genetic ancestry.
